# Implementation and Evaluation of the Harmonized Cognitive Assessment Protocol in an Arabic-Speaking Population: The Lebanon Study on Aging and Health (LSAHA)

**DOI:** 10.64898/2026.09.02.26362058

**Authors:** Martine Elbejjani, Khalil El Asmar, Monique Chaaya, Ghia Sanjar, Aya El Sammak, Kenneth M Langa, Stephen J McCall, Abla Sibai, Carlos Mendes De Leon

## Abstract

The Middle East and North Africa (MENA) region is projected to experience the fastest growth in dementia burden globally, yet population-based data on cognitive aging remain scarce. We present the implementation and psychometric evaluation of a comprehensive cognitive assessment in the Lebanon Study on Aging and Health (LSAHA), the first large-scale, population-based study of cognitive aging and dementia in an Arab country.

We implemented a construct-centered adaptation of the Harmonized Cognitive Assessment Protocol (HCAP), integrating culturally-relevant and field-informed adaptation and implementation procedures. The battery was administered in a large, socioeconomically and educationally diverse sample of older adults in Lebanon (n=3,027). Confirmatory factor analyses (CFA) were used to evaluate psychometric performance.

CFA results demonstrated good to excellent model fit across general and domain-specific cognition. Cognitive scores showed expected trajectories across age, education, and subjective health and memory indicators. Findings were consistent with HCAP studies across multiple countries, supporting cross-national comparability.

This study establishes a foundational infrastructure for cognitive aging research in Lebanon and the broader Middle-East and North Africa (MENA) region and demonstrates the feasibility and value of adapting HCAP for underrepresented populations.

## 1. INTRODUCTION

Estimates from the Global Burden of Disease Study indicate that the Middle-East and North Africa (MENA) region will experience the steepest increases in dementia burden globally by 2050.^1^ Yet, Arab countries continue to face major gaps in evidence on Alzheimer’s disease and related dementias (ADRD) prevalence, burden, and determinants, leaving them under-prepared to address this growing challenge.^2^ Lebanon, a lower-middle-income country (LMIC), is one of the fastest-aging countries in the region and is projected to experience among the largest increases in ADRD rates.^1, 3^ This rapid population aging is occurring amid chronic and overlapping crises. Older adults in Lebanon have experienced substantial lifecourse adversities, including a 15-year civil war (1975-1990), chronic political instability, and recurrent episodes of violence. These exposures have been compounded by severe crises in later life, including one of the world’s most severe socioeconomic collapses, the 2020 Beirut Port blast, and renewed war exposure.^4, 5^ Existing data, though sparse, also suggest considerable cardiovascular and metabolic burdens.^3, 6^ Together, these lifecourse and health exposures may confer a heightened vulnerability to ADRD that remains largely unexamined, limiting understanding of distinctive risk pathways and hindering public health planning.

A fundamental barrier to advancing cognitive aging research in Lebanon and the MENA region is the limited availability of validated cognitive assessments for Arabic-speaking older adults. A systematic review conducted by our group underscored these limitations, identifying only 20 validated cognitive tools, with just three validated in more than one setting/study (the Mini-Mental State Examination (MMSE), Montreal Cognitive Assessment (MoCA), and Addenbrooke’s Cognitive Examination III (ACE III)). Beyond tool scarcity, the review revealed major gaps in psychometric evidence and a lack of evaluations of domain-specific assessments and cognitive constructs. Existing studies predominantly rely on small samples and brief screening tools that lack the depth required to evaluate cognitive function across domains, detect early impairment, and model cognitive aging trajectories.^2^

To address these gaps, the Lebanon Study on Aging and Health (LSAHA) was launched as the first large-scale population-based study of cognitive aging and dementia in an Arab country. LSAHA incorporated a cognitive battery based on the Harmonized Cognitive Assessment Protocol (HCAP), a flexible yet standardized framework designed to produce robust and internationally comparable cognitive data.^7^ HCAP has been implemented in leading aging studies in over 19 countries, including the US, England, Indica, China, Mexico, and South Africa, with additional planned studies around the world.^8^ LSAHA represents the first HCAP-aligned implementation in an Arabic-speaking population. In this study, we describe the development and adaptation of the LSAHA-HCAP cognitive battery and evaluate its psychometric performance using confirmatory factor analysis. These efforts provide foundational data to advance local population-level evidence on cognitive aging and enable cross-national research with data from an understudied world region.

## 2. MATERIALS AND METHODS

### 2.1. Study design and sample

LSAHA enrolled participants aged ≥60 years from two regions: Beirut (the capital) and the Zahle district (Bekaa governate) to capture Lebanon’s urban-rural, socioeconomic, and religious diversity. The study employed a probability-based multi-stage sampling design to recruit 1,500 participants per site. Sampling procedures have been described previously.^9^ Briefly, all residential areas in Beirut and 25 villages/towns in Zahle were selected using probability proportional to population size, and divided into 100×100 to 400×400 meters grids. Grids were proportionally allocated within these areas and a random sample of grids was selected for recruitment. Household enumeration was conducted within selected grids to identify age-eligible adults. Up to two adults aged ≥60 years were selected per household; in households with more than two age-eligible adults, two were randomly selected. Eligibility criteria were being ≥60 years, ability to speak Arabic, residing in the household for at least three months in the past year, and having no plans to leave Lebanon within six months. Recruitment and data collection started on October 9, 2023 and was completed on September 10, 2024.

The study was approved by the American University of Beirut Institutional Review Board. All participants provided written informed consent following a brief decision-making competency assessment; individuals who did not pass the assessment (n=159) were not enrolled.

### 2.2. Data collection

Data collection involved a computer-assisted face-to-face standardized interview administered by trained interviewers at the participant’s home. The two-hour interview included cognitive assessments and standardized measures of demographic, social, economic, and health indicators, including lifecourse socioeconomic indicators, retirement, physical and mental health, and contextual exposures such as war/conflict exposure, economic hardships, and healthcare access. Anthropometric and physical assessments, blood samples, and a key informant interview were also collected.^9^

### 2.3. Cognitive assessments

#### 2.3.1. Cognitive battery development

The LSAHA cognitive battery was developed through a construct-centered adaptation of the HCAP, prioritizing appropriateness for the cultural, linguistic, and literacy diversity of the older population in Lebanon while ensuring harmonization with international HCAP studies. To guide battery development, given the limited availability of cognitive tools in our context, we conducted a systematic review of validated assessments among Arabic-speaking older adults ^2^, confirming the 10/66 Dementia Research Group protocol,^10^ previously implemented in Lebanon with good psychometric performance,^11^ as the strongest available foundation.^2^ The 10/66 battery includes items from the Community Screening Instrument for Dementia (CSI-D)^12^ and the Consortium to Establish a Registry of Alzheimer’s Disease (CERAD).^11, 13^ Items from the 10/66 battery were mapped to four cognitive domains (orientation, language, memory, and visuospatial). As the 10/66 battery lacked executive function assessments, we incorporated the Symbol Digit Modalities Test (SDMT),^14^ the Symbol Cancellation Test (SCT),^15^ and two similarities items from the MoCA.^16^ Selection of these additions prioritized adaptation feasibility, performance in prior HCAP studies,^17^ use in local clinical settings,^18^ and available translation and validity documentation (e.g., the MoCA emerged in our review as one of the most used instruments in Lebanon and the region).^2^ We also expanded the memory domain by adding a delayed story recall and the language domain to include a full FAS verbal fluency test (by adding two additional letter tests to the original 10/66).^19^ All 10/66 and added items were reviewed with local geriatricians and HCAP consultants for conceptual and practical relevance.

The resulting LSAHA-HCAP battery was a 30-45 minutes assessment of five domains (Table 1): Orientation was assessed using ten items, including orientation to time and place and two contextual orientation items (current president and a historical figure/event). Memory tests included the CERAD ^13^ immediate (sum of three trials) and delayed word recall, immediate and delayed story recall (“Brave Man”), and the CSI-D 3-word immediate and delayed recall and delayed name recall.^11, 12^ Visuospatial ability assessment included two CERAD constructional praxis items (circle, pentagon).^13^ Language/fluency assessments included the animal naming test, the FAS verbal fluency test, and the CSI-D object naming, object describing, repeat a phrase, instruction (nod, point), and three-step instruction items. Executive function was assessed using the SCT, SDMT, and two MoCA similarities items (ruler/watch, bicycle/train).

**Table 1.**
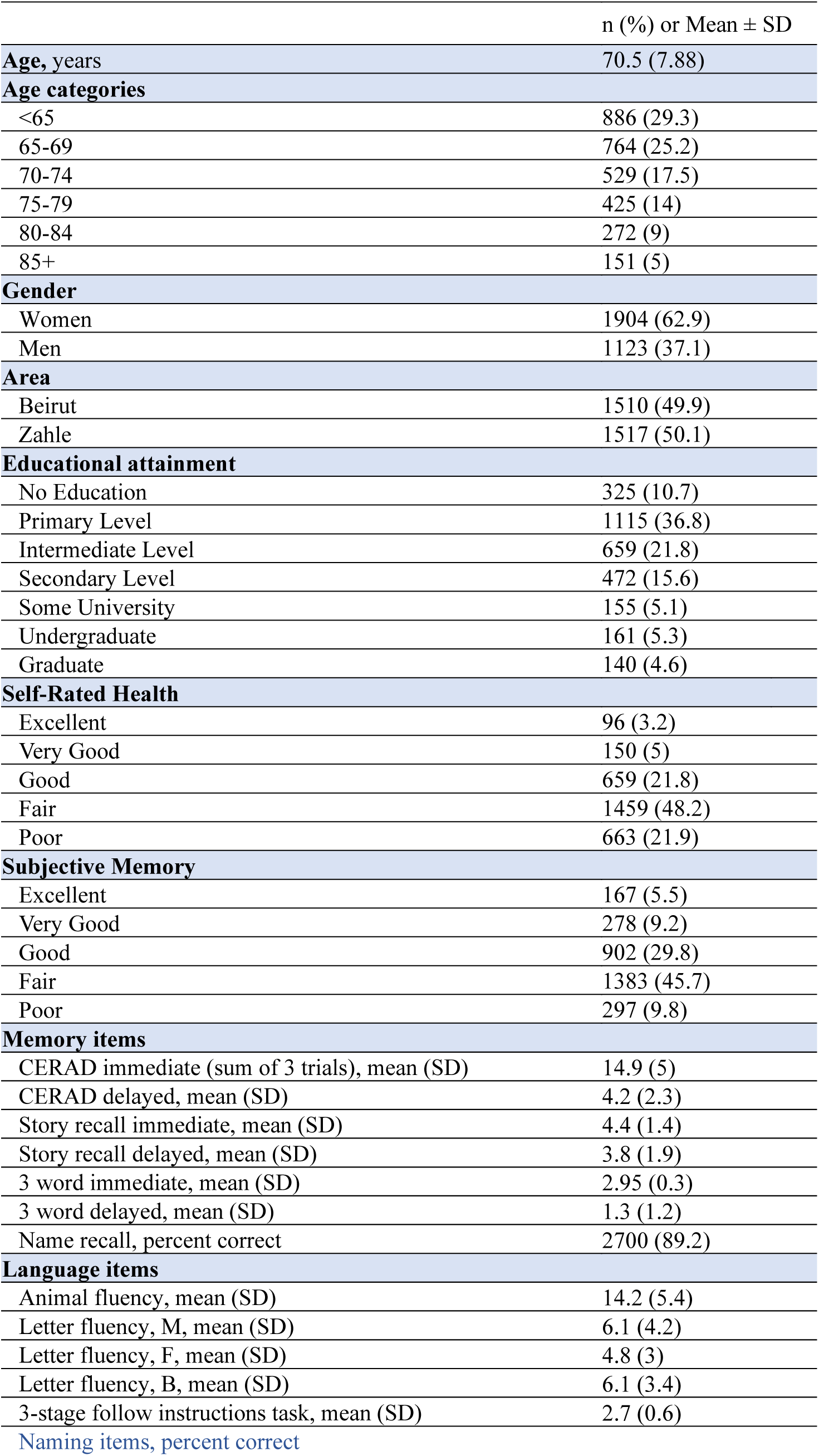
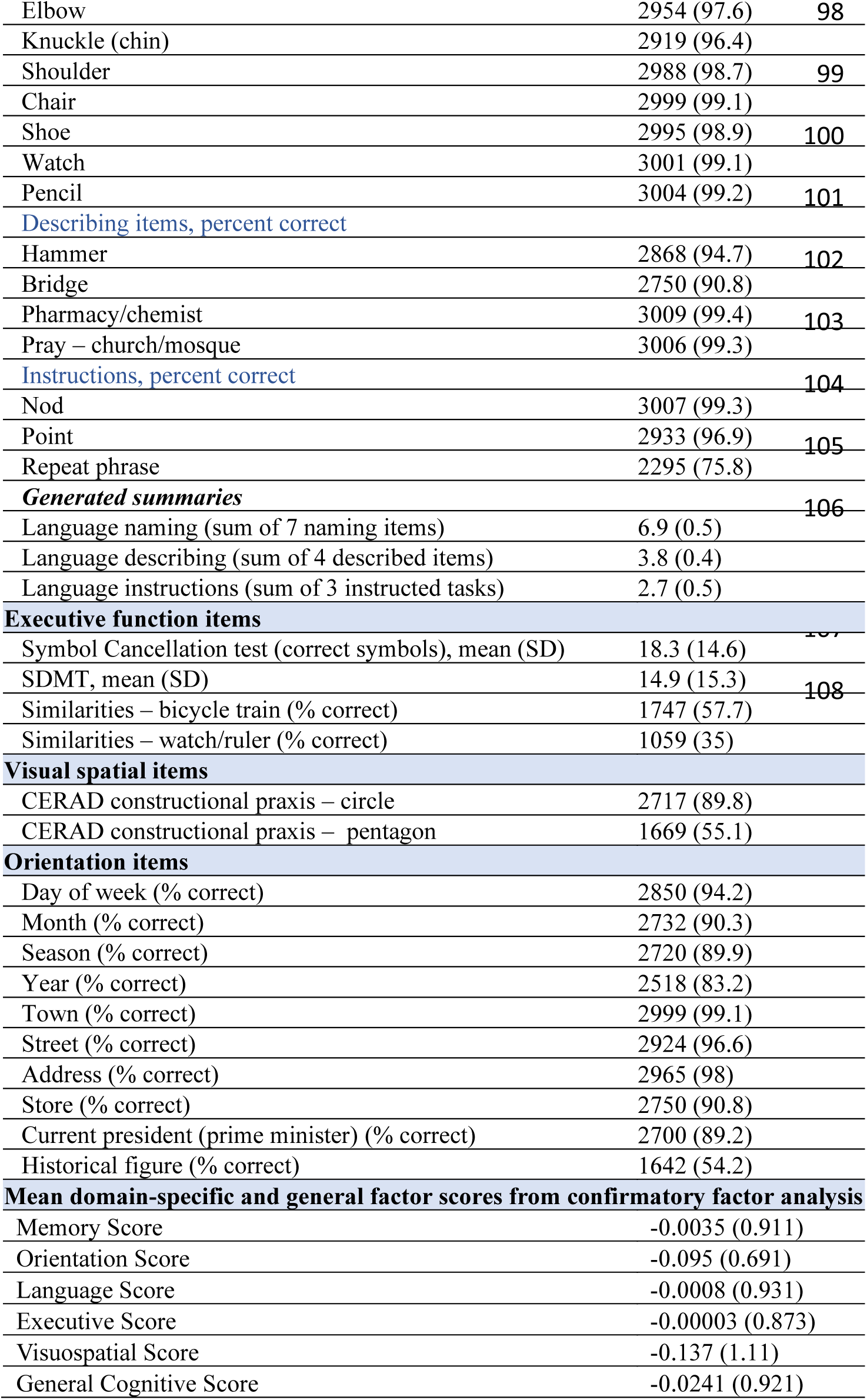
Descriptive characteristics of the LSAHA sample (N=3,027)

| | n (%) or Mean ± SD |
| --- | --- |
| **Age, years** | 70.5 (7.88) |
| **Age categories** |  |
| <65 | 886 (29.3) |
| 65-69 | 764 (25.2) |
| 70-74 | 529 (17.5) |
| 75-79 | 425 (14) |
| 80-84 | 272 (9) |
| 85+ | 151 (5) |
| **Gender** |  |
| Women | 1904 (62.9) |
| Men | 1123 (37.1) |
| **Area** |  |
| Beirut | 1510 (49.9) |
| Zahle | 1517 (50.1) |
| **Educational attainment** |  |
| No Education | 325 (10.7) |
| Primary Level | 1115 (36.8) |
| Intermediate Level | 659 (21.8) |
| Secondary Level | 472 (15.6) |
| Some University | 155 (5.1) |
| Undergraduate | 161 (5.3) |
| Graduate | 140 (4.6) |
| **Self-Rated Health** |  |
| Excellent | 96 (3.2) |
| Very Good | 150 (5) |
| Good | 659 (21.8) |
| Fair | 1459 (48.2) |
| Poor | 663 (21.9) |
| **Subjective Memory** |  |
| Excellent | 167 (5.5) |
| Very Good | 278 (9.2) |
| Good | 902 (29.8) |
| Fair | 1383 (45.7) |
| Poor | 297 (9.8) |
| **Memory items** |  |
| CERAD immediate (sum of 3 trials), mean (SD) | 14.9 (5) |
| CERAD delayed, mean (SD) | 4.2 (2.3) |
| Story recall immediate, mean (SD) | 4.4 (1.4) |
| Story recall delayed, mean (SD) | 3.8 (1.9) |
| 3 word immediate, mean (SD) | 2.95 (0.3) |
| 3 word delayed, mean (SD) | 1.3 (1.2) |
| Name recall, percent correct | 2700 (89.2) |
| **Language items** |  |
| Animal fluency, mean (SD) | 14.2 (5.4) |
| Letter fluency, M, mean (SD) | 6.1 (4.2) |
| Letter fluency, F, mean (SD) | 4.8 (3) |
| Letter fluency, B, mean (SD) | 6.1 (3.4) |
| 3-stage follow instructions task, mean (SD) | 2.7 (0.6) |
| Naming items, percent correct |  |

|  |  |  |
| --- | --- | --- |
| Elbow | 2954 (97.6) | 98 |
| Knuckle (chin) | 2919 (96.4) |  |
| Shoulder | 2988 (98.7) | 99 |
| Chair | 2999 (99.1) |  |
| Shoe | 2995 (98.9) | 100 |
| Watch | 3001 (99.1) |  |
| Pencil | 3004 (99.2) | 101 |
| *Describing items, percent correct* |  |  |
| Hammer | 2868 (94.7) | 102 |
| Bridge | 2750 (90.8) |  |
| Pharmacy/chemist | 3009 (99.4) | 103 |
| Pray – church/mosque | 3006 (99.3) |  |
| *Instructions, percent correct* |  |  |
| Nod | 3007 (99.3) | 104 |
| Point | 2933 (96.9) | 105 |
| Repeat phrase | 2295 (75.8) |  |
| *Generated summaries* |  |  |
| Language naming (sum of 7 naming items) | 6.9 (0.5) | 106 |
| Language describing (sum of 4 described items) | 3.8 (0.4) |  |
| Language instructions (sum of 3 instructed tasks) | 2.7 (0.5) |  |
| **Executive function items** |  |  |
| Symbol Cancellation test (correct symbols), mean (SD) | 18.3 (14.6) | 107 |
| SDMT, mean (SD) | 14.9 (15.3) | 108 |
| Similarities – bicycle train (% correct) | 1747 (57.7) |  |
| Similarities – watch/ruler (% correct) | 1059 (35) |  |
| **Visual spatial items** |  |  |
| CERAD constructional praxis – circle | 2717 (89.8) |  |
| CERAD constructional praxis – pentagon | 1669 (55.1) |  |
| **Orientation items** |  |  |
| Day of week (% correct) | 2850 (94.2) |  |
| Month (% correct) | 2732 (90.3) |  |
| Season (% correct) | 2720 (89.9) |  |
| Year (% correct) | 2518 (83.2) |  |
| Town (% correct) | 2999 (99.1) |  |
| Street (% correct) | 2924 (96.6) |  |
| Address (% correct) | 2965 (98) |  |
| Store (% correct) | 2750 (90.8) |  |
| Current president (prime minister) (% correct) | 2700 (89.2) |  |
| Historical figure (% correct) | 1642 (54.2) |  |
| **Mean domain-specific and general factor scores from confirmatory factor analysis** |  |  |
| Memory Score | -0.0035 (0.911) |  |
| Orientation Score | -0.095 (0.691) |  |
| Language Score | -0.0008 (0.931) |  |
| Executive Score | -0.00003 (0.873) |  |
| Visuospatial Score | -0.137 (1.11) |  |
| General Cognitive Score | -0.0241 (0.921) |  |

#### 2.3.2. Adaptation & Implementation

Battery items were reviewed with local geriatricians and field coordinators of the earlier 10/66 study for cultural, translation, and linguistic appropriateness and field applicability, with input from HCAP consultants on feasibility and administration across diverse populations. Items and tests were further evaluated through cognitive interviewing with older adults.^20^ Adaptations included substituting formal words in the word recall and naming/describing lists with familiar dialect alternatives, adapting orientation items related to location to allow landmark-based responses, due to Lebanon’s lack of standardized addresses systems, and adapting FAS letters based on a linguistic review of letters in Arabic with difficulty and frequency equivalent to F, A, and S, supported by a review of prior FAS and MoCA Arabic adaptations and local clinical experience. We also retained modifications implemented in the original 10/66 adaptation (Supplementary Material).

We developed standardized administration and scoring manuals and interviewers completed an eight-day training program with supervised practice, scoring calibration, and field-readiness assessments. We implemented multi-level quality assurance procedures including field observations, recordings review, and analytical data checks. Through these procedures, we identified an alternative response to the historical figure/event item that was close in timing and context and we coded both responses as correct. We also found inconsistencies in the SCT incorrect symbols scoring, and, although total SCT scores and correct-symbol scores were strongly correlated, we opted to only use the latter in our analysis to minimize measurement error. Data monitoring helped understand low scores on the watch/ruler item which frequently reflected a nuanced conceptual distinction, as in the local context, a watch is associated with “giving time” rather than “measuring time”; the item was retained given good model fit indices.

### 2.4. Statistical Analysis

Following HCAP conventions, “don’t know” responses were coded as 0 and “refused” as missing. Overall, 638 participants (21%) had at least one missing cognitive item/test; item/test-level missingness was <11.7% and was <3% for the majority of test/items. We imputed missing observations using a k-nearest neighbor imputation (KNN) and a dataset with broad background indicators, including health, demographic, socioeconomic, and lifestyle variables. We conducted our analyses using the imputed data and confirmed results were concordant in the unimputed dataset.

We summarized sample characteristics and cognitive items using means and proportions. Continuous cognitive items were scaled prior to model estimation to improve numerical stability, and binary and ordinal indicators were modeled as ordered categorical variables. We fit a series of unidimensional and hierarchical confirmatory factor analysis (CFA) models, starting with baseline domain-specific models and a full hierarchical model without any initial modifications. These models demonstrated good (baseline orientation and hierarchical model), acceptable (memory and language), and poor (executive function) fit (Supplementary Table S1). Model fit for all domains was subsequently improved through a series of theoretically and empirically informed modifications.^21–25^ For the memory domain, we added residual covariances between the immediate and delayed story recall and between the immediate and delayed word recall tasks, consistent with model improvements documented in previous HCAP studies.^21–24^ For executive function, we added a residual correlation between the two similarities items. For language, which included several binary items with little variance (Table 1), we improved model fit by using sum scores for sets of binary-scored items: object naming (range 0–7), description (0–4), and following instruction items (0–3). Finally, the historical event/figure item, which is conceptually mapped as “long-term memory” in the 10/66 was reassigned to the orientation domain, based on theoretical rationale (contextual orientation), improved model performance (including better factor loadings and fit indices (Supplementary Table S2)), and stronger correlations with orientation items. The visuospatial domain had only two items and was specified as a just-identified model (df = 0), for which global fit indices are necessarily perfect.^21,23^

Once adequate fit was obtained for all unidimensional models, we combined them into a hierarchical multiple domain factor analysis estimating general cognitive function scores. CFA model fit was ascertained using three standard fit statistics: the Root Mean Square Error of Approximation (RMSEA), Comparative Fit Index (CFI), and Standardized Root Mean Residual (SRMR), applying standard HCAP thresholds.^21, 23, 24, 26^ Internal consistency reliability was assessed using Cronbach’s alpha; the visuospatial domain included only two indicators and was therefore evaluated using the inter-item association rather than a conventional multi-item alpha.

We further evaluated the construct validity of estimated domain-specific and general cognitive factors by correlating their z-scores with demographic characteristics (age, gender, and education); we also explored their association with self-rated health and subjective memory (“How would you rate your memory at the present time?”), using linear regressions adjusting for age, sex, education, and study site. Analyses were conducted using R (R Core Team, 2024).

## 3. RESULTS

The LSAHA participants (n=3027) were on average 70.5 years old (SD=7.88; range=60–98 years) and 63% were female (Table 1); 10.7% had no education, 36.8% had primary education, and 9.9 % had university-level education. Descriptive statistics for raw cognitive items/tests and factor scores are presented in Table 1.

### 3.1. Confirmatory factor analyses

After implementing model modifications, domain-specific and general cognition CFA models showed good to excellent fit (RMSEA≤0.05 and CFI≥95); SRMR for language and general cognition models were slightly over 0.05 and SRMR for orientation was around 0.08, indicating satisfactory fit.^23^ Memory and executive function models showed the best fit (Table 2).

**Table 2.** Fit statistics for unidimensional and hierarchical confirmatory analysis models in the LSAHA sample (N=3,027)

|  | # of tests | RMSEA | CFI | SRMR | Implemented model specifications |
| --- | --- | --- | --- | --- | --- |
| Memory | 7 | 0.014 | 1.00 | 0.032 | *Correlated items: immediate~delayed story & Immediate~delayed 10-word recall* |
| Language | 8 | 0.046 | 0.980 | 0.058 | *Generating summary scores for:*<br>*-naming items (pencil, watch, shoes, chair, knuckle, shoulder, elbow)*<br>*-describing items (bridge, hammer, chemist, church/mosque)*<br>*-binary instruction items (nod, point, repeat phrase)* |
| Executive Function | 4 | 0.000 | 1.00 | 0.006 | *Correlated items: similarities* |
| Orientation | 10 | 0.023 | 0.983 | 0.082 |  |
| Visuospatial | 2 | 0 | 1 | 0 | *Just identified model* |
| General cognition |  | 0.026 | 0.988 | 0.058 |  |

RMSEA= Root Mean Square Error of Approximation; CFI= Comparative Fit Index; SRMR= Standardized Root Mean Residual (SRMR). Model fit was considered excellent if $CFI=1$ and $RMSEA=0$ and $SRMR=0$ , good if $CFI \geq 0.95$ and $RMSEA \leq 0.05$ and $SRMR \leq 0.05$ , adequate if $CFI \geq 0.90$ and $RMSEA \leq 0.08$ and $SRMR \leq 0.08$ , and poor if either $CFI < 0.9$ or $RMSEA > 0.08$ or $SRMR > 0.08$ .

The final hierarchical CFA model is presented in Figure 1. Factor loadings were within good and acceptable ranges (0.4-0.9). The lowest loadings were observed in the language domain for following instructions (0.345), the 3-stage task (0 .376), and object naming (0 .385), likely reflecting limited variability and ceiling effects. Factor loadings in unidimensional models were generally similar to those in the hierarchical model (Supplementary Table S3). Internal consistency was high (alpha=0.8-0.9) for most domains (memory=0.90, language=0.79, executive function=0.70, orientation=0.88, and visuospatial=0.86). Model structure and fit were comparable using the un-imputed data (Supplementary Tables S4-S5 and Supplementary Figure S1).

**Fig 1.**
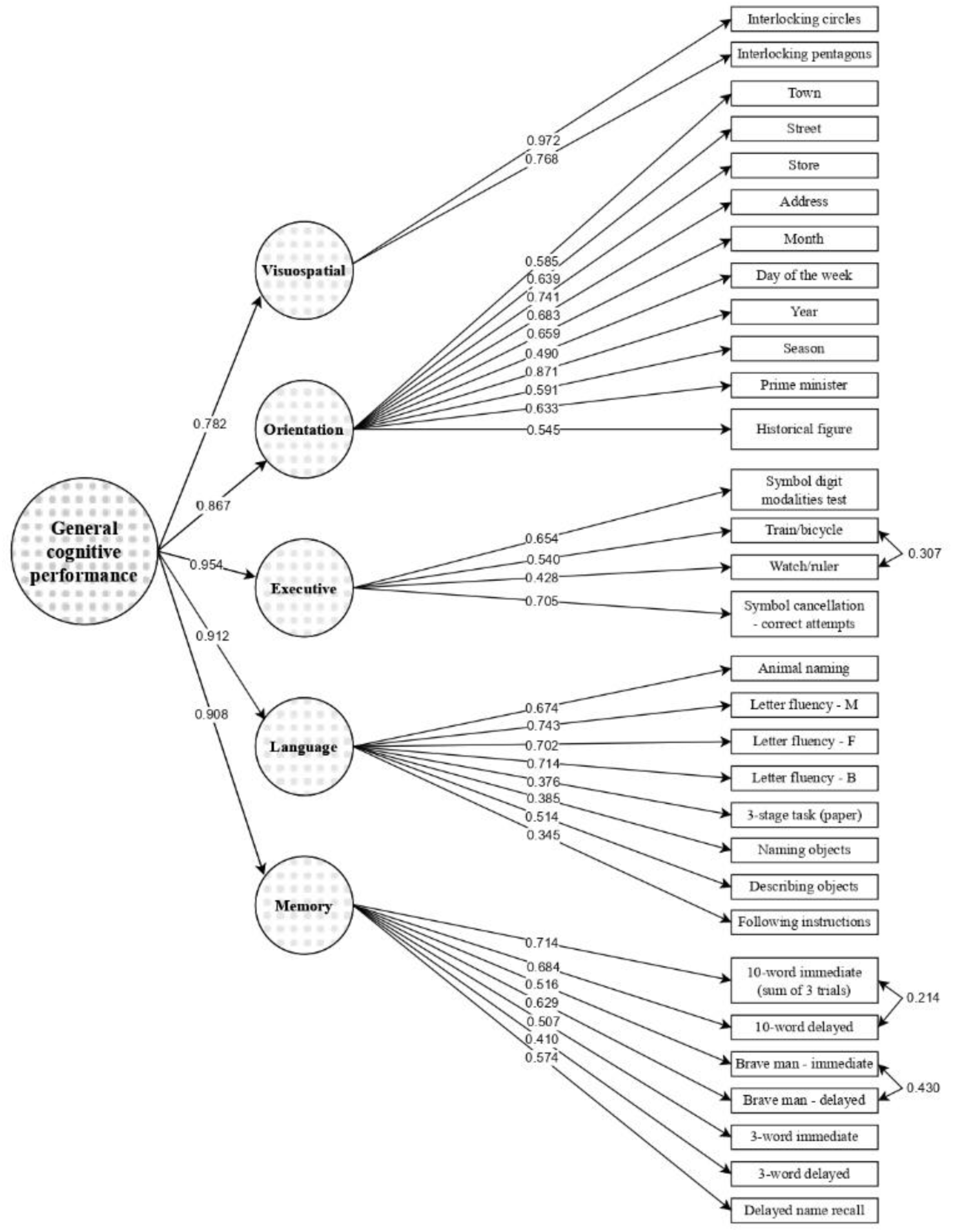
Model diagram of the hierarchical confirmatory factor analysis in the LSAHA sample (N=3,027) Diagram shows standardized factor loadings; circles represent latent variables for general and domain-specific cognition, boxes represent the sets of cognitive tests forming latent variables. Double-headed arrows show residual correlations between item/test scores.

General and domain-specific factor scores were relatively normally distributed (Supplementary Figure S2), while distribution of orientation scores was skewed, reflecting ceiling effects, as observed in similar HCAP analyses.^24, 26^ Correlations between domain-specific factor scores ranged from 0.40 to 0.60, with strongest correlations (∼0.60) observed between memory, language, and executive function and lowest correlations (∼0.40) observed between visuospatial scores and other domains (Supplementary Figure S3).

#### Cognitive factor scores across sociodemographic indicators and self-rated health and memory

General and domain-specific cognitive scores were negatively correlated with age, with strongest correlations observed for general cognition (-0.41), memory (-0.41), and executive function (-0.38; Figure 2A). Cognitive scores were positively correlated with education, with stronger correlations observed for general cognition, executive function, and language scores (>0.5), and markedly lower scores for participants with no education (Figure 2B). These patterns were similar in men and women, although men had higher cognitive scores (except for memory) and correlations with age and education were stronger among women (Supplementary Figure S4). We further examined these correlations across lower (<secondary) and higher (≥secondary) education strata, given gender-differences in educational attainment in our sample (Supplementary Figure S5). Men and women with higher educational attainment had comparable cognitive scores across age groups (Figure 3). However, in the lower education group, women had lower cognitive scores (except for memory), with a trend for increasing disparities with increasing age, particularly for general cognition, language, orientation, and visuospatial scores. Adjusted cognitive scores were consistently lower among participants reporting poorer self-rated health and subjective memory (Supplementary Figure S6).

**Fig 2.**
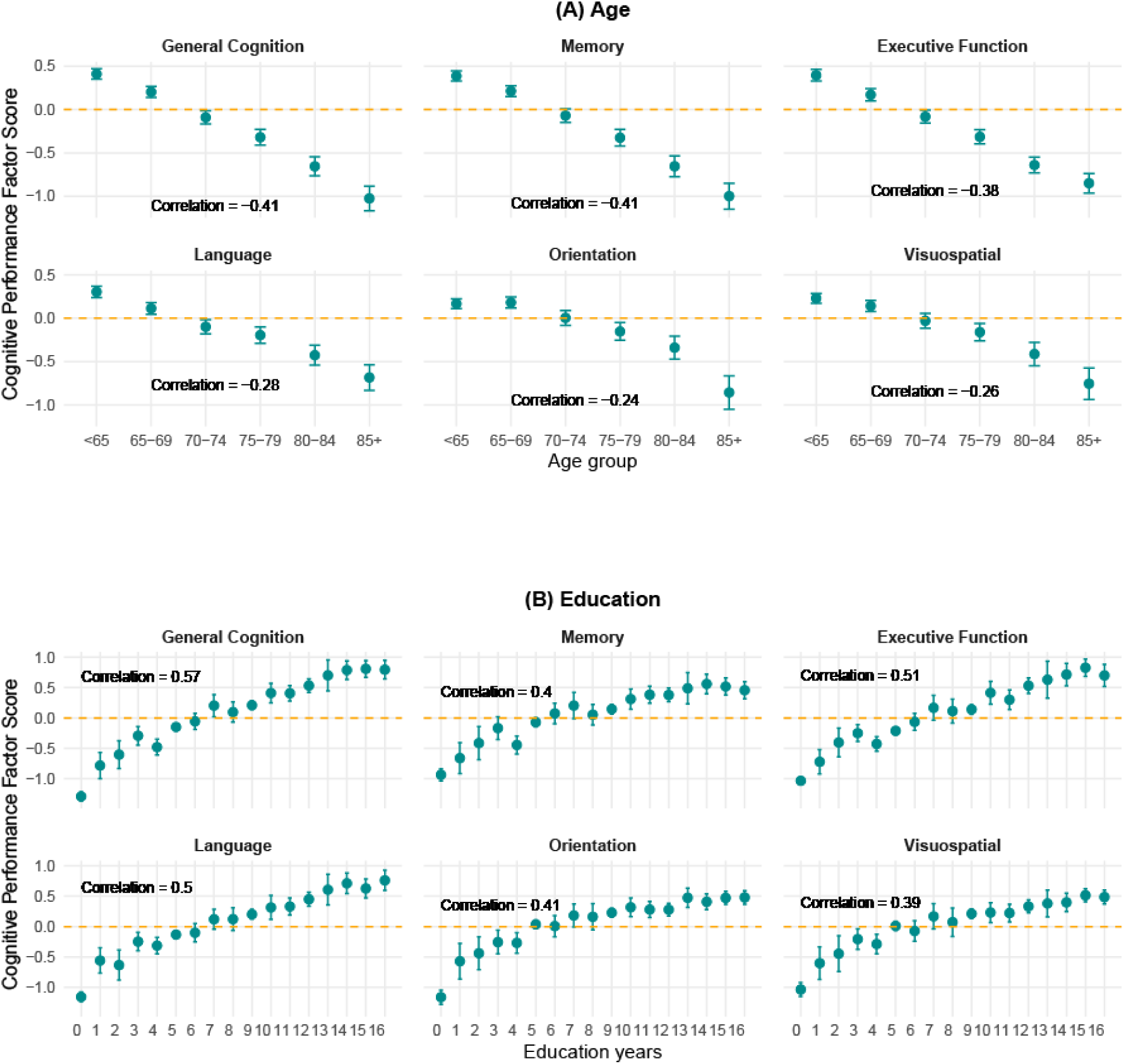
Mean general and domain−specific cognitive scores by age (A) and education (B) levels in the LSAHA sample (N = 3,027)

**Fig 3.**
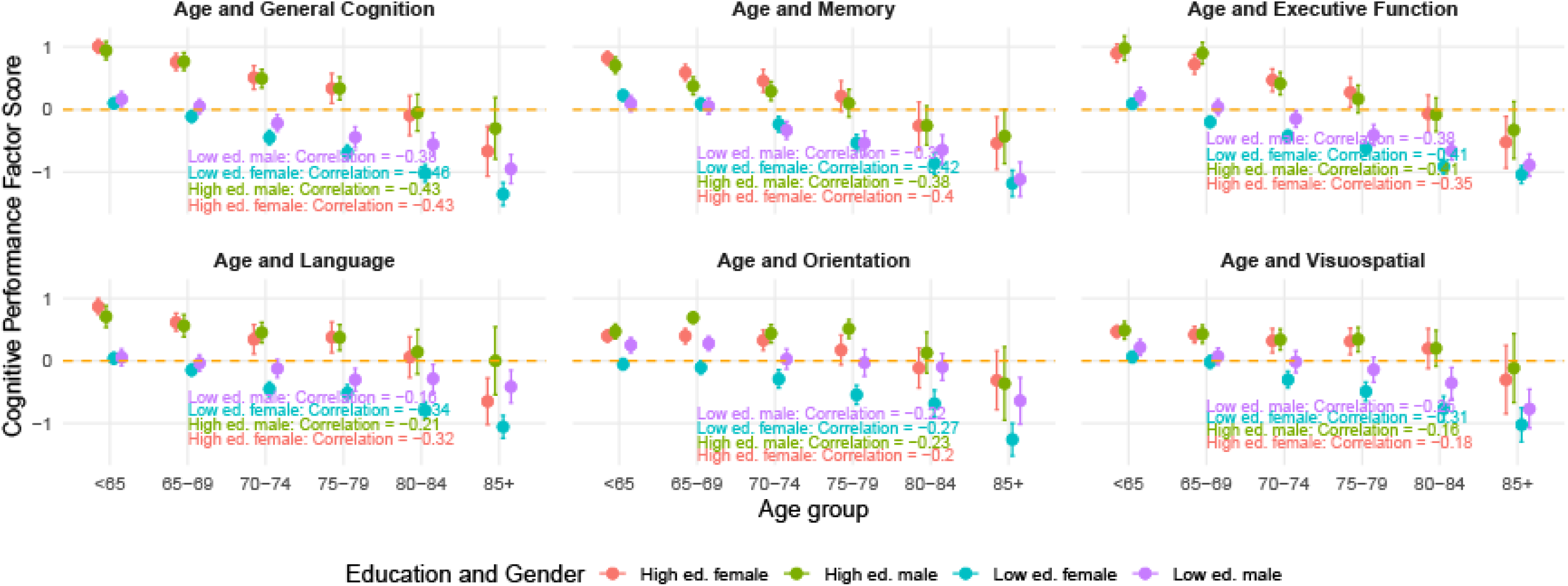
Mean general and domain-specific cognitive scores by gender and education groups in the LSAHA sample (N=3,027)

## 4. DISCUSSION

This study presents the first implementation and evaluation of a comprehensive cognitive battery in a population-based sample of older adults in Lebanon. The LSAHA-HCAP battery, developed through a construct-centered adaptation of the HCAP, was tailored to the Lebanese context and administered to a large socioeconomically diverse sample. Confirmatory factor analyses showed that the battery captured general and domain-specific cognitive function with good to excellent model fit, and cognitive scores tracked coherently with age, education, self-rated health, and subjective memory. Results were highly consistent with those of HCAP studies across the world, including the US, England, India, Mexico, China, South Africa, and Kenya,^21–24, 26, 27^ supporting the cross-cultural applicability of the HCAP framework. In a context of limited validated tools and psychometric evidence, our findings demonstrate the value and feasibility of implementing adapted HCAP assessments in the MENA region and understudied LMICs settings. LSAHA provides a foundation for advancing evidence on cognitive aging in Lebanon and for evaluating unique contextual risk factors, including lifecourse exposure to conflict and resource insecurities. The study also expands the breadth of harmonized cognitive aging research and cross-country comparisons into an underrepresented world region.

Domain-specific and general cognition CFA models showed good to excellent fit, with memory and executive function having the best performance. Language and orientation models showed relatively lower fit, although still within good (RMSEA and CFI) and satisfactory (SRMR) thresholds, likely due to limited variability in some items. For example, in the language domain, simpler tasks (e.g., object naming, describing, and instructions) with lower variability had lower loadings than animal naming and FAS letter tests, which potentially capture the fluency spectrum better. Overall, fit indices were comparable if not more favorable than other HCAP studies, though slightly weaker for language.^21, 23, 24, 26^

Several elements likely contributed to the robust performance of the LSAHA-HCAP battery. First, battery design, test selection, and implementation incorporated best practices and resourceful lessons drawn from other HCAP studies,^28^ particularly those in LMIC and global south settings, such as India (LASI-DAD), South Africa (HAALSI), and Mexico (Mex-Cog).^21, 29, 30^ Second, our study benefitted from prior field implementation of the 10/66 protocol in Lebanon and incorporated insight from local clinical settings and cognitive interviewing.^11, 20^ Analytically, we applied model refinements shown to improve model fit in HCAP studies.^21–23^ The strong performance of the executive function domain is particularly notable. This domain was designed for the LSAHA-HCAP battery, as it was not part of the 10/66 protocol. Despite the lack of psychometric evidence on executive function assessments among Arabic-speaking older adults,^2^ this domain showed excellent fit and expected correlations with other domains, age, education, and subjective health and memory. These results support the feasibility of extending domain coverage, using adaptable tests and items from established instruments (the MoCA in our case). Overall, results highlight the value of context-sensitive adaptation and the expanding collective capacity of the HCAP network, to which this study contributed further insights from an understudied population with limited psychometric evidence.^28^

Factor loadings generally fell within acceptable ranges (0.4-0.9) and were comparable to previous HCAP studies. Items with lower loadings (∼0.4), such as the 3-word immediate recall, instruction, and object naming items were similarly found to have weaker loadings in other HCAP studies,^21, 24, 26^ reflecting ceiling effects and limited variability common to these items, particularly in general populations. Nonetheless, their conceptual value and comparability across studies together with good model fit indices justify retaining them and the additional performance information they may carry. Distributions of domain-specific and general cognitive scores and correlations among domains were also comparable to HCAP results from several samples.^21, 24, 26^

General and domain-specific cognitive scores followed expected patterns across age and education and were markedly lower among participants with poorer self-rated health and memory. General cognition, memory, and executive function scores showed the strongest negative correlations with age, consistent with other HCAP analyses.^24, 26^ Gender disparities were evident, with women showing lower cognitive scores across all domains. One exception was the trend for higher memory scores among women, in line with results from the US, England, India, and Mexico HCAP studies.^26^ Gender differences were most pronounced at lower educational levels, suggesting a cumulative disadvantage, while scores were comparable between men and women in the higher education group. Women were also found to have lower cognitive scores in India (LASI-DAD), Mexico (Mex-Cog), China (CHARLS-HCAP), and South Africa (HAALSI-HCAP); in contrast, they had higher or comparable cognitive scores in the US HRS-HCAP and the English ELSA-HCAP, respectively.^26^ Together, these findings underscore persistent and widespread gender disparities in cognition across various LMICs. Our results further indicate that this disadvantage was predominantly among women with lower educational attainment and that disparities worsened in older age groups. Future LSAHA analyses will elucidate the interplay between age, education, gender, and cohort effects, and the role of lifecourse psychosocial and cardiovascular/metabolic risk in shaping cognitive outcomes and gender disparities. Another important future direction will be the generation of normative cognitive scores and algorithm classifications of mild cognitive impairment and ADRD.

### 4.1. Strengths and limitations

This first population-based implementation of an HCAP-aligned battery in Lebanon demonstrated robust psychometric performance and consistency with international HCAP findings. Key strengths include a construct-centered HCAP adaptation, cultural adaptation and implementation informed by local field and clinical experience and by HCAP best practices and lessons, and rigorous quality monitoring in a large and diverse sample.

The study also has several limitations. First, although the sampling strategy aimed to capture socioeconomic and urban-rural diversity, the sample is not randomly drawn from the entire population of older adults in Lebanon, due to logistical and security challenges, limiting generalizability. Second, due to consent and ethical considerations, individuals with more severe cognitive or functional impairment may be under-represented, limiting our findings and battery performance towards healthier ranges of cognitive abilities. Third, ceiling effects in some orientation and language items may reduce sensitivity in detecting subtle cognitive differences in these domains. Fourth, administration and interpretation challenges may have increased measurement error in certain items. For example, the watch/ruler item responses could reflect conceptual differences in perceiving the question rather than incorrect answers. We also opted to accept responses for the historical figure item that were contextually close to the correct response, and we used the SCT-correct symbol score, rather than the full tallied score. While these issues were limited to a few items and overall model fit remained strong, they may have introduced additional measurement noise and could affect comparability for those specific measures. Lastly, the cross-sectional design limits the ability to disentangle aging processes from cohort differences in education, socioeconomic opportunities, and health profiles; longitudinal follow-up data will be essential to capture within-person aging-related cognitive changes.

### 4.2. Conclusion

The LSAHA-HCAP battery demonstrated strong psychometric performance and captured meaningful variation in cognitive function across age, education, gender, and health indicators in a large socioeconomically diverse sample of older adults in Lebanon. The alignment of our findings with HCAP studies across high-, middle-, and low-income countries underlines the value of harmonized cognitive assessments. By extending harmonized cognitive aging research into the MENA region, LSAHA establishes a critical foundation for investigating contextual determinants and contributes to global efforts to address rising dementia burden in historically underrepresented and vulnerable populations.

## ACKNOWLEDGEMENTS

The authors would like to thank the LSAHA participants for their invaluable time and positive engagement with the study. We extend our gratitude to the HCAP network for their continued support. We would also like to acknowledge the study data collectors as well as the municipalities and community members for their support.

## DATA AVAILABILITY STATEMENT

Study details and protocols are available on the study website (https://sites.aub.edu.lb/lsaha/).

Deidentified data are available by applying for data requests.

## CONSENT STATEMENT

The study was approved by the Institutional Review Board of the American University of Beirut. All participants provided written informed consent.

## Author contributions

**ME:** conceptualization, data curation, formal analysis, funding acquisition, investigation, methodology, project administration, resources, software, supervision, validation, visualization, writing – original draft, and writing– review & editing. **KEA:** data curation, formal analysis, methodology, validation, and writing– review & editing. **MC:** conceptualization, data curation, funding acquisition, investigation, project administration, resources, and writing– review & editing. **GS:** data curation, formal analysis, methodology, validation, and writing– review & editing. **AES**: data curation, formal analysis, investigation, methodology, project administration, software, and writing– review & editing. **KML**: conceptualization, investigation, methodology, and writing– review & editing. **SJM**: funding acquisition, investigation, and writing– review & editing. **AM**: conceptualization, funding acquisition, investigation, and writing– review & editing. **CML**: conceptualization, data curation, formal analysis, funding acquisition, investigation, methodology, project administration, resources, software, supervision, and writing– review & editing.

## SUPPORTING IFORMATION CAPTIONS

## Tables

Supplementary Table S1. Fit statistics for baseline unidimensional and hierarchical confirmatory analysis models (without modifications, using initial items available for each domain) in the LSAHA sample (N=3,027)

Supplementary Table S2. Fit statistics for the memory and orientation domain with different mapping of the historical figure item in the LSAHA sample (N=3,027)

Supplementary Table S3. Factor loadings from unidimensional confirmatory factor analysis in the LSAHA sample (N=3,027)

Supplementary Table S4. LSAHA sample characteristics and missingness in imputed and un-imputed datasets (N=3,027)

Supplementary Table S5. Fit statistics for final unidimensional and hierarchical confirmatory analysis models in the LSAHA sample (using unimputed dataset with complete observations, N=2,389)

## Figures

Supplementary Figure S1. Model diagram of the hierarchical confirmatory factor analysis using original un-imputed data of the LSAHA study (N=2,389)

Supplementary Figure S2. Distribution of general and domain-specific factor scores derived from confirmatory factor analysis in the LSAHA study (N=3,027)

Supplementary Figure S3. Correlation of domain-specific factor scores derived from confirmatory factor analyses in the LSAHA sample (N=3,027)

Supplementary Figure S4. Mean general cognitive and domain-specific cognitive scores by age (A) and education levels (B) among male and female participants in the LSAHA sample (N=3,027)

Supplementary Figure S5. Proportion of male and female participants with lower educational attainment across age groups in the LSAHA sample (N=3,027)

Supplementary Figure S6. Adjusted mean general and domain−specific cognitive scores across self−rated health (A) and subjective memory (B) ratings

