## Supplementary Material for "Implementation and Evaluation of the Harmonized Cognitive Assessment Protocol in an Arabic-Speaking Population: The Lebanon Study on Aging and Health (LSAHA)"

### Details on LSAHA-HCAP item/test adaptations

- **Memory - CERAD word recall and CSI-D 3-word word recall lists & Language - naming and describing object lists:**

We substituted formal words with familiar dialect-appropriate alternatives (e.g., hand, boat, and hammer); we replaced knuckle with chin as there is no word for knuckle in Arabic.

- **Orientation items:**

Given the lack of standardized addresses systems in Lebanon, responses were adapted to accept responses based on nearby landmarks, major streets, or commonly known reference location points.

"Prime Minister/Name of Mayor" was replaced with the name of current president of Lebanon.

The CSI-D long-term memory/orientation question (What is the name of the civil rights leader who was assassinated in Memphis in 1968?) was replaced with a contextual equivalent (Who was the president that was assassinated after three weeks in office in 1982 during the first Israeli invasion of Lebanon?). *Some responses included a historical assassination close in context and timing (1989) and we considered both responses as correct.

- **Language domain - FAS letter adaptations:**

The FAS letters were adapted to FMB based on a linguistic review of letters in Arabic with difficulty and frequency equivalent to F, A, and S; this step was complemented by a review of prior FAS and MoCA adaptations in Arabic as well as insights from practitioners' experience in administering these tests in local clinical settings.

- **Language domain - CSI-D repeat a phrase:**

Adapting the repeat phrase to a Lebanese tongue twister.

**Supplementary Table S1. Fit statistics for baseline unidimensional and hierarchical confirmatory analysis models (without modifications, using initial items available for each domain) in the LSAHA sample (N=3,027)**

| | # of Items/tests | RMSEA | CFI | SRMR |
|---|---|---|---|---|
| Memory | 8 | 0.103 | 0.961 | 0.091 |
| Language | 19 | 0.051 | 0.906 | 0.163 |
| Executive Function | 4 | 0.201 | 0.882 | 0.113 |
| Orientation | 9 | 0.021 | 0.985 | 0.082 |
| Visuospatial | 2 (Just identified model) | 0 | 1 | 0 |
| **General cognition baseline model** | | 0.042 | 0.949 | 0.122 |

RMSEA= Root Mean Square Error of Approximation; CFI= Comparative Fit Index; SRMR= Standardized Root Mean Residual (SRMR).

**Supplementary Table S2. Fit statistics for the memory and orientation domain with different mapping of the historical figure item in the LSAHA sample (N=3,027)**

| | # of Items/tests | RMSEA | CFI | SRMR |
|---|---|---|---|---|
| Memory Unidimensional Model – with the historical figure item | 8 | 0.029 | 0.995 | 0.037 |
| Memory Unidimensional Model – without the historical figure item | 7 | 0.014 | 1.00 | 0.032 |
| Orientation Unidimensional Model – with the historical figure item | 10 | 0.023 | 0.983 | 0.082 |
| Orientation Unidimensional Model – without the historical figure item | 9 | 0.021 | 0.985 | 0.082 |
| Full Hierarchical Model | | | | |
| General cognition – historical figure item in the memory domain | | 0.029 | 0.986 | 0.060 |
| General cognition – historical figure item in orientation domain | | 0.028 | 0.986 | 0.060 |

RMSEA= Root Mean Square Error of Approximation; CFI= Comparative Fit Index; SRMR= Standardized Root Mean Residual (SRMR).

**Supplementary Table S3. Factor loadings from unidimensional confirmatory factor analysis in the LSAHA sample (N=3,027)**

| | |
|---|---|
| **Memory items** | |
| CERAD immediate (sum of 3 trials) | 0.696 |
| CERAD delayed | 0.716 |
| Story recall immediate | 0.506 |
| Story recall delayed | 0.615 |
| 3 word immediate | 0.461 |
| 3 word delayed | 0.492 |
| Name recall | 0.518 |
| **Language items** | |
| Animal fluency | 0.605 |
| Letter fluency, M | 0.780 |
| Letter fluency, F | 0.802 |
| Letter fluency, B | 0.793 |
| 3-stage follow instructions task, mean (SD) | 0.306 |
| Language naming (sum of 7 naming items) | 0.365 |
| Language describing (sum of 4 described items) | 0.411 |
| Language instructions (sum of 3 instructed tasks) | 0.292 |
| **Executive function items** | |
| Symbol Cancellation test (correct symbols) | 0.772 |
| SDMT | 0.777 |
| Similarities – bicycle train | 0.418 |
| Similarities – watch/ruler | 0.265 |
| **Visual spatial items** | |
| CERAD constructional praxis – circle | 0.864 |
| CERAD constructional praxis – pentagon | 0.864 |
| **Orientation items** | |
| Day of week | 0.529 |
| Month | 0.685 |
| Season | 0.778 |
| Year | 0.599 |
| Town | 0.643 |
| Street | 0.678 |
| Address | 0.707 |
| Store | 0.724 |
| Current president (prime minister) | 0.653 |
| Historical figure | 0.564 |

**Supplementary Table S4. LSAHA sample characteristics and missingness in imputed and un-imputed datasets (N=3,027)**

| | **Un-Imputed**<br>n (%) or Mean ± SD | ***n missing*** | **Imputed**<br>n (%) or Mean ± SD |
|---|---|---|---|
| **Age,** years | 70.5 (7.88) | | 70.5 (7.88) |
| **Age categories** | | | |
| <65 | 886 (29.3) | | 886 (29.3) |
| 65-69 | 764 (25.2) | | 764 (25.2) |
| 70-74 | 529 (17.5) | | 529 (17.5) |
| 75-79 | 425 (14) | | 425 (14) |
| 80-84 | 272 (9) | | 272 (9) |
| 85+ | 151 (5) | | 151 (5) |
| **Gender** | | | |
| Women | 1904 (62.9) | | 1904 (62.9) |
| Men | 1123 (37.1) | | 1123 (37.1) |
| **Area** | | | |
| Beirut | 1510 (49.9) | | 1510 (49.9) |
| Zahle | 1517 (50.1) | | 1517 (50.1) |
| **Educational attainment** | | 18 (0.6) | |
| No Education | 324 (10.7) | | 325 (10.7) |
| Primary Level | 1106 (36.5) | | 1115 (36.8) |
| Intermediate Level | 652 (21.5) | | 659 (21.8) |
| Secondary Level | 471 (15.6) | | 472 (15.6) |
| Some University | 155 (5.1) | | 155 (5.1) |
| Undergraduate | 161 (5.3) | | 161 (5.3) |
| Graduate | 140 (4.6) | | 140 (4.6) |
| **SRH** | | 120 (4) | |
| Excellent | 96 (3.2) | | 96 (3.2) |
| Very Good | 150 (5) | | 150 (5) |
| Good | 657 (21.7) | | 659 (21.8) |
| Fair | 1409 (46.5) | | 1459 (48.2) |
| Poor | 595 (19.7) | | 663 (21.9) |
| **Subjective Memory** | | 124 (4.1) | |
| Excellent | 167 (5.5) | s | 167 (5.5) |
| Very Good | 278 (9.2) | | 278 (9.2) |
| Good | 891 (29.4) | | 902 (29.8) |
| Fair | 1278 (42.2) | | 1383 (45.7) |
| Poor | 289 (9.5) | | 297 (9.8) |
| **Cognitive factor scores from domain-specific and hierarchical confirmatory factor analysis models** | | | |
| Memory Scores | -0.00426 (0.911) | 309 | -0.00349 (0.911) |
| Orientation Scores | -0.095 (0.691) | 32 | -0.095 (0.691) |
| Language Scores | -0.00211 (0.925) | 211 | -0.00075 (0.931) |
| Executive Scores | -0.0000608 (0.875) | 412 | -0.0000271 (0.873) |
| Visuospatial Scores | -0.138 (1.1) | 105 | -0.137 (1.11) |
| General Cog Score | -0.0254 (0.920) | 638 | -0.0241 (0.921) |

**Supplementary Table S5. Fit statistics for final unidimensional and hierarchical confirmatory analysis models in the LSAHA sample (using unimputed dataset with complete observations, N=2,389)**

| | # of Items/tests | RMSEA | CFI | SRMR |
|---|---|---|---|---|
| Memory | 8 | 0.023 | 0.999 | 0.042 |
| Language | 19 | 0.099 | 0.923 | 0.068 |
| Executive Function | 4 | 0.009 | 1 | 0.009 |
| Orientation | 9 | 0.023 | 0.983 | 0.082 |
| Visuospatial | 2 (Just identified model) | 0 | 1 | 0 |
| **Full model** | | 0.038 | 0.978 | 0.068 |

RMSEA= Root Mean Square Error of Approximation; CFI= Comparative Fit Index; SRMR= Standardized Root Mean Residual (SRMR).

**Supplementary Figure S1**. **Model diagram of the hierarchical confirmatory factor analysis using original un-imputed data of the LSAHA study (N=2,389)**

*Diagram shows standardized factor loadings; circles represent latent variables for general and domain-specific cognition, boxes represent the sets of cognitive tests forming latent variables. Double-headed arrows show residual correlations between item/test scores.*

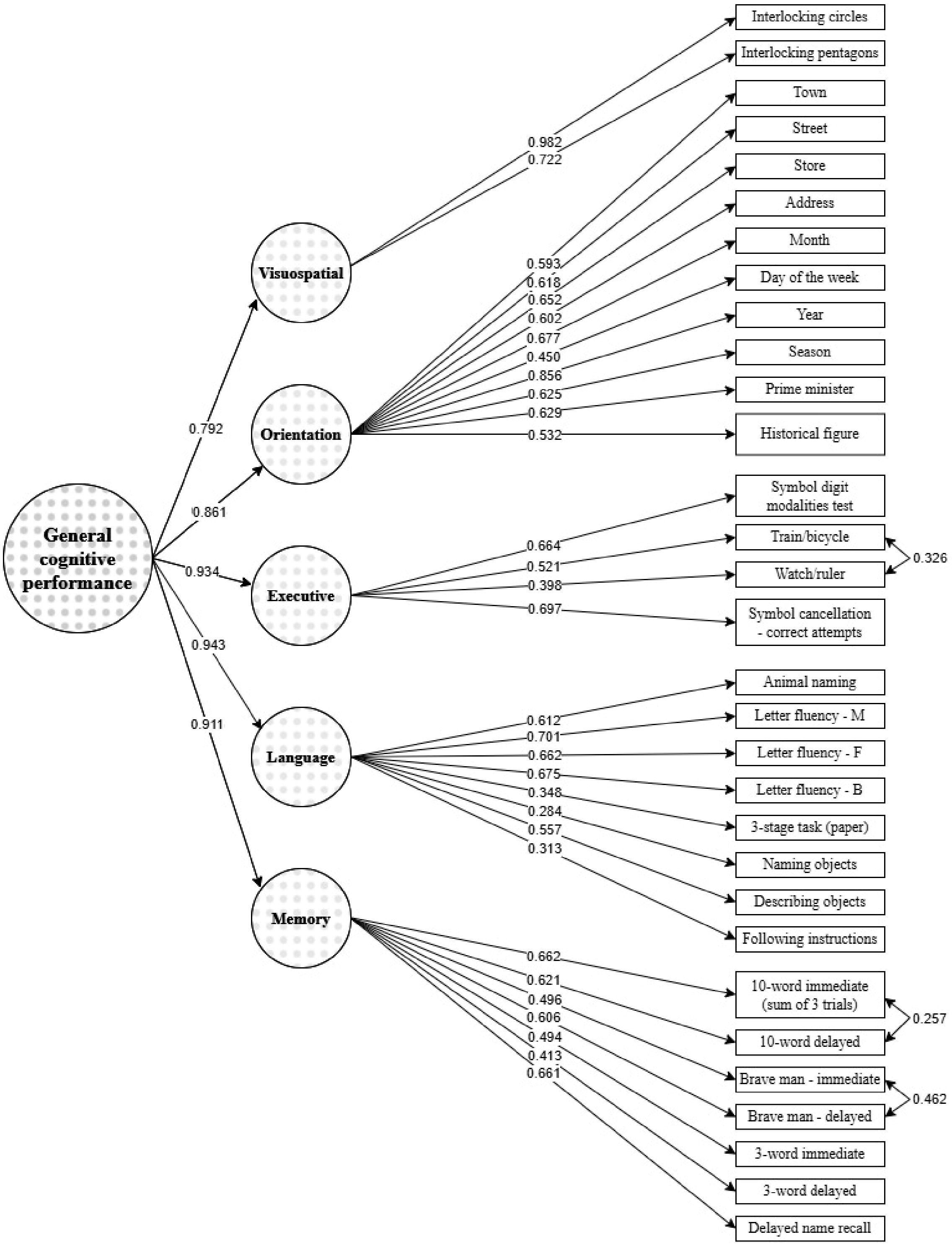

**Supplementary Figure S2. Distribution of general and domain-specific factor scores derived from confirmatory factor analysis in the LSAHA study (N=3,027)**

*Distribution of z-scores is presented. The y-axis scale is different to follow the distribution range and shape of each score*

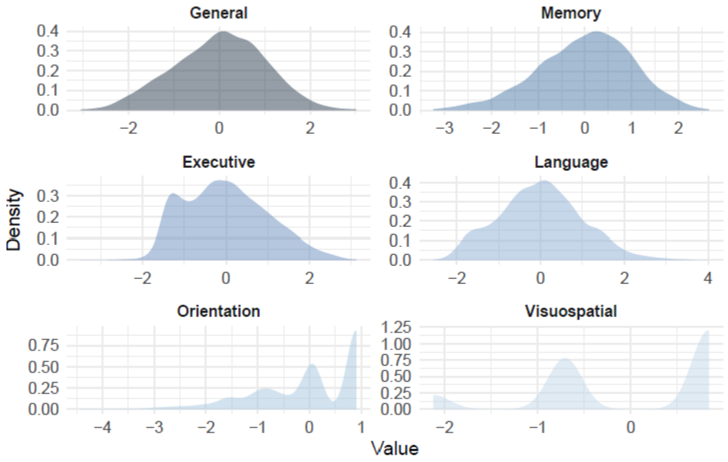

**Supplementary Figure S3. Correlation of domain-specific factor scores derived from confirmatory factor analyses in the LSAHA sample (N=3,027)**

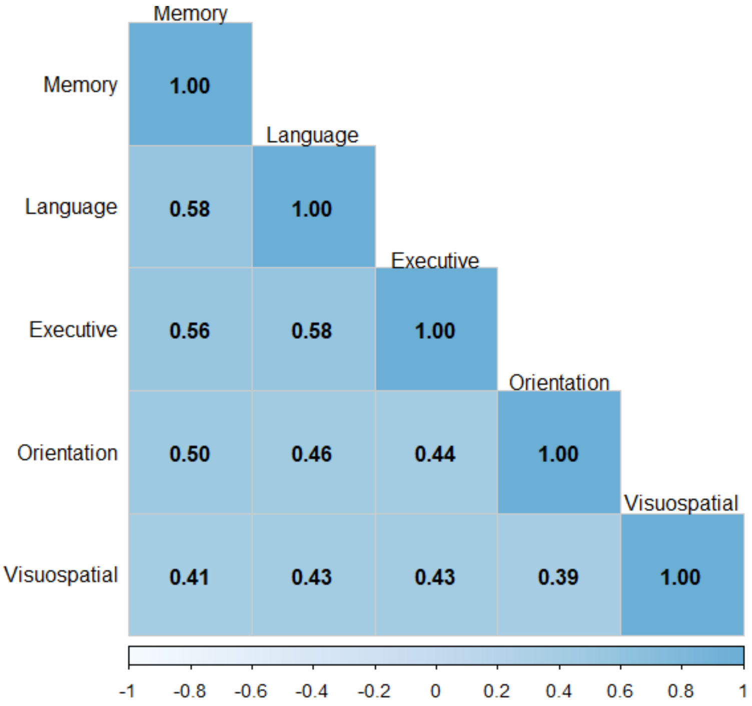

**Supplementary Figure S4. Mean general cognitive and domain-specific cognitive scores by age (A) and education levels (B) among male and female participants in the LSAHA sample (N=3,027)**

**A.**

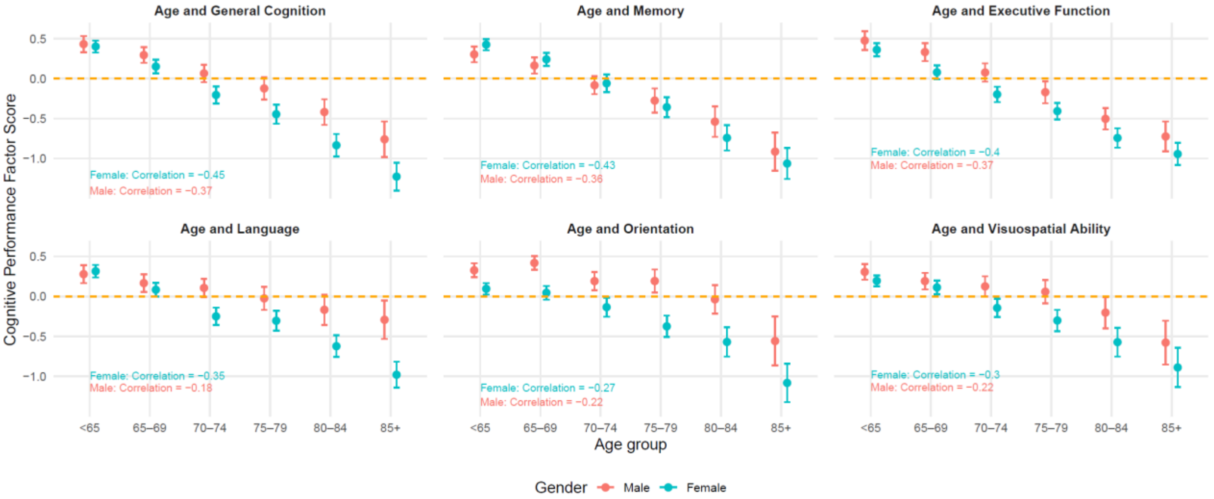

**B.**

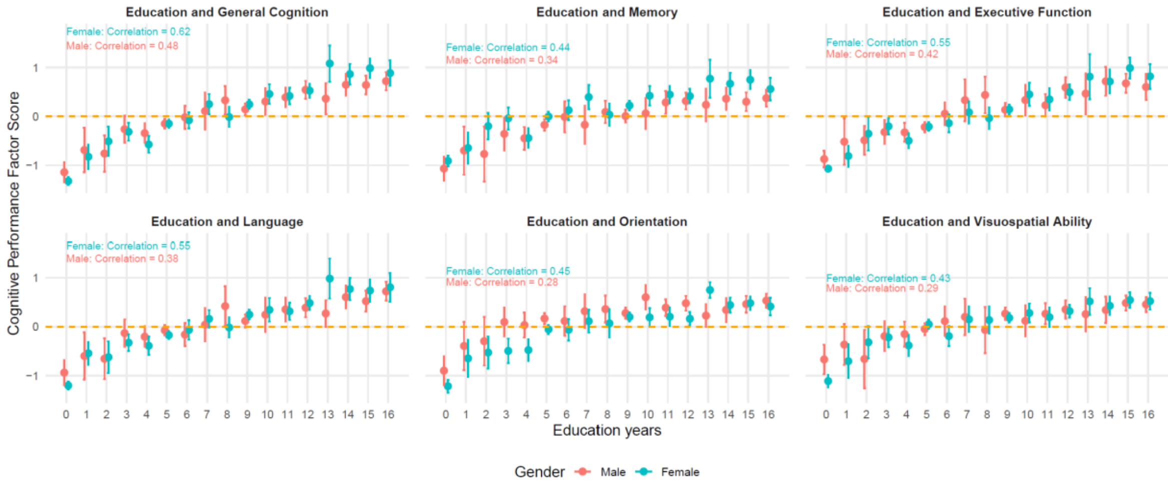

**Supplementary Figure S5. Proportion of male and female participants with lower educational attainment across age groups in the LSAHA sample (N=3,027)**

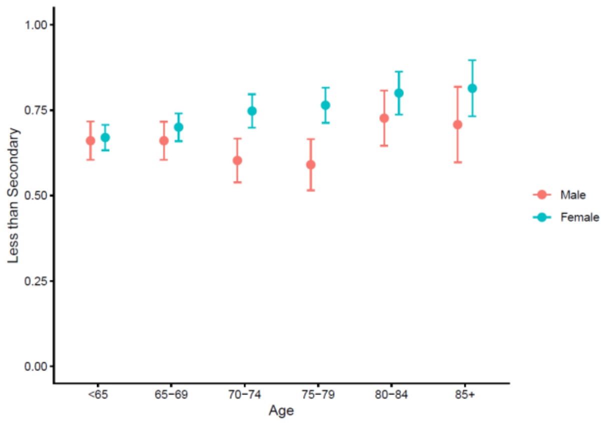

**Supplementary Figure S6. Adjusted mean general and domain−specific cognitive scores across self−rated health (A) and subjective memory (B) ratings**

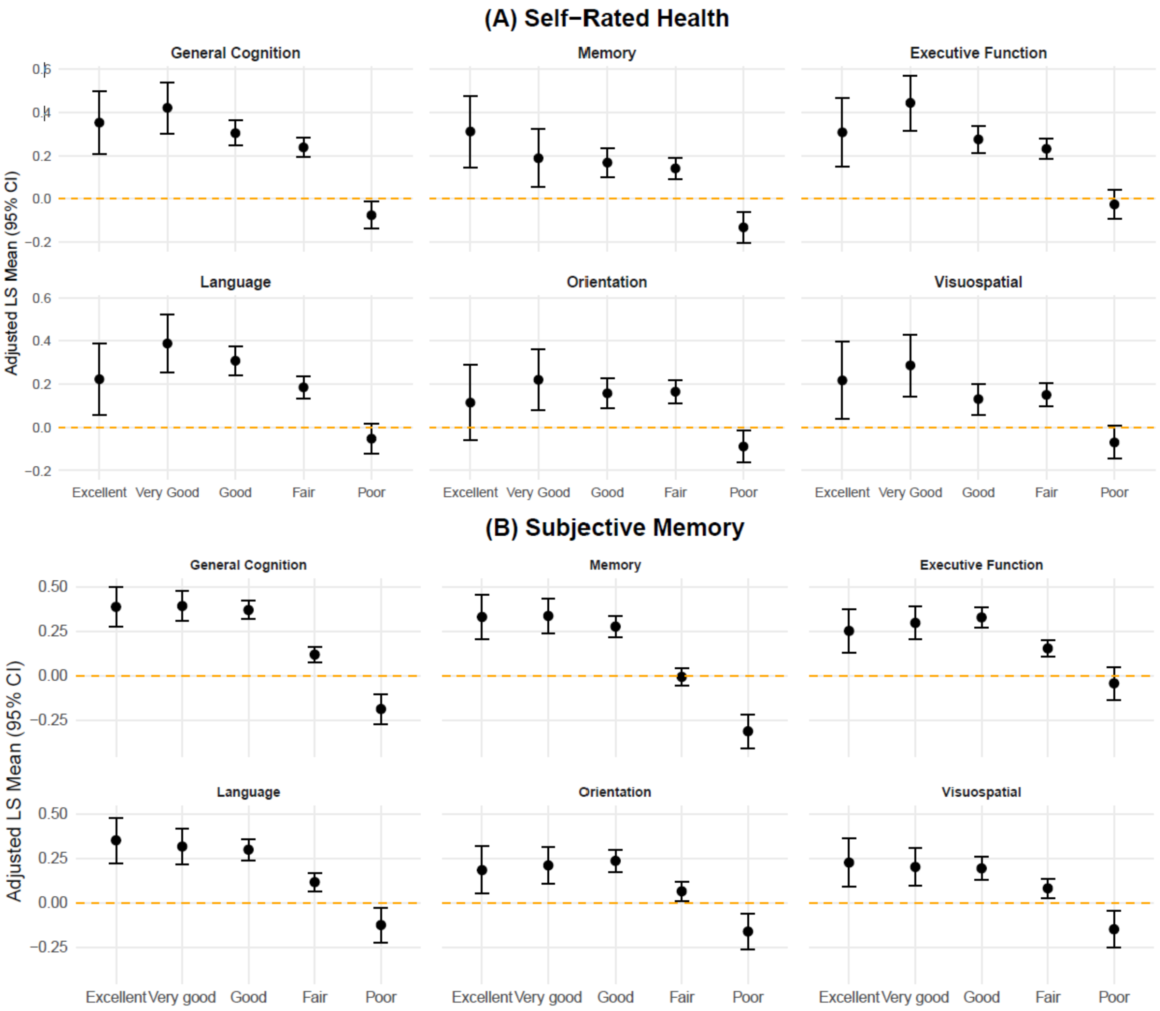
